# Vaccine Prioritisation Using Bluetooth Exposure Notification Apps

**DOI:** 10.1101/2020.12.14.20248186

**Authors:** Mark D Penney, Yigit Yargic, Lee Smolin, Edward W Thommes, Madhur Anand, Chris T. Bauch

## Abstract

After vaccinating health care workers and vulnerable groups against COVID-19, authorities will need to decide how to vaccinate everyone else. Prioritising individuals with more contacts can be disproportionately effective, in theory, but identifying these individuals is difficult. Here we show that the technology underlying Bluetooth exposure notification applications, such as used for digital contact tracing, can be leveraged to prioritise vaccination based on individual contact data. Our approach is based on the insight that these apps also act as local sensing devices measuring each user’s total exposure time to other users, thereby enabling the implementation of a previously impossible strategy that prioritises potential super-spreaders. Furthermore, by generalising percolation theory and introducing a novel measure of vaccination efficiency, we demonstrate that this “hot-spotting” strategy can achieve herd immunity with up to half as many vaccines as a non-targeted strategy, and is attractive even for relatively low rates of app usage.

---

Since the first recorded cases in late 2019, COVID-19 has spread like wildfire across the globe. In order to contain the spread of the disease, an array of non-pharmaceutical interventions have been deployed, including hygienic measures, isolation of infected individuals, quarantine of their exposed individuals, social distancing, and even large scale closures of businesses and institutions [41]. This stimulated efforts to develop COVID-19 vaccines in record time [24]. In late 2020, the first phase of COVID-19 vaccination began with a small number of health-care workers and elderly individuals receiving the Pfizer/BioNTech vaccine. These groups were recommended for vaccine prioritisation [4, 3] based on the high rate of mortality and other severe outcomes experienced by elderly individuals [13] and the principle of reciprocity, which states that those who accepted the greatest risks to mitigate the effects of the pandemic, should be vaccinated first [36].

Once the most vulnerable individuals have been vaccinated, public health authorities will need to decide how to vaccinate the rest of the population[27, 21, 12]. In 2021, data on whether available COVID-19 vaccines prevent both transmissibility and disease in vaccinated individuals will become increasingly available [18]. Hence, public health authorities may want to consider pivoting to a strategy that interrupts transmission of SARS-CoV-2 most effectively. And in all likelihood, the supply of vaccines and/or the capacity to administer them will continue to be limited in early 2021[24], necessitating additional prioritisation decisions.

In this work we study how to prioritise COVID-19 vaccines in the second phase, where vaccination of vulnerable groups has been achieved. We assume a goal of achieving herd immunity through interrupting transmission as efficiently as possible, and we explore a scenario where COVID-19 vaccines can prevent transmissibility as well as disease, as applies to many other existing vaccines against respiratory pathogens [10].

Prioritisation of vaccines is often based on age or other demographic factors, and we know that contact patterns differ between demographic groups. However, as demonstrated in empirical studies of contact patterns [30], heterogeneity within demographic groups is much greater than the differences between them. Because individuals with more contacts have the potential to fuel the spread more than others, prioritising individuals for vaccination according to the risk they pose to further spread has been found to be highly effective in previous theoretical models [35, 19, 16, 14]. However, such exposure prioritisation strategies are difficult to implement in practice since public health authorities only have information on broad demographic or regional factors upon which to act.

It is exactly in this regard that Bluetooth exposure notification apps have opened up a new avenue for public health interventions. The core functionality of these apps is the creation of an encounter log between app users. This encounter log is useful for more than just exposure notification: It also quantifies the user’s exposure to others. Said another way, these apps are also sensors which measure the duration of exposure: an epidemiologically significant quantity.

Though other digital contact tracing technologies exist, Bluetooth-based solutions have been chosen when the protection of user’s personally identifiable information was a core concern. The data in the encounter log can only be used by public health authorities in a manner which does not require the centralized collection of user data. In fact, in our proposal each user’s device makes a decision *purely locally*, based on the number of encounters reported, as to whether or not they will be prioritised for vaccination. By preferentially vaccinating individuals with greater total exposure to others our proposed strategy is significantly more efficient than traditional prioritisation strategies.

The success of digital contact tracing technologies has been hampered by inadequate uptake rates [23, 11]. Our proposal doesn’t suffer from this issue. We show that the efficiency depends only on the fraction of the app-using population which receive the vaccine, with the greatest relative reduction occurring with the vaccination of roughly 20-40% of app users.

Our modelling approach is based on percolation theory: a collection of analytical techniques, coming from statistical physics [39], which has been successfully applied to material sciences [37] and to the spread of forest fires [25, 26] and infectious diseases [17]. Here we extend the theory in two directions: Firstly, we introduce a novel measure to compare vaccine prioritisation strategies; and secondly, we incorporate heterogeneity in the risk of infection (total exposure time) between contacts. These tools allow us to show that the improved efficiency gained by piggy-backing vaccine prioritisation strategies on top of exposure notification apps is a robust phenomenon, as it derives its power from the very heterogeneities in contact patterns that shape the spread of COVID-19.

## Bluetooth Exposure Notification

In March 2020, COVID Watch released a white paper detailing an anonymous Bluetooth-based system that exploits the ubiquity of Android and iOS smartphones to support contact tracing [20]. The idea has seen widespread adoption, with nearly every developed country having incorporated it into their digital contact tracing solutions. Both the Google/Apple and BlueTrace frameworks are also based on this technology, the former of which is available throughout North America and the European Union and the latter in Singapore and Australia.

Protecting users’ privacy was a fundamental principle underlying the design process. Every 10-20 minutes anonymous tokens are exchanged between app users who are in close proximity to one another. Each user’s device stores these tokens in an encounter log. The data stored by the app is able to determine neither the number of contacts nor the duration of any individual contact. Instead, the number of tokens logged over a given timeframe is a measure of the user’s total exposure time to others: the sum, over each contact, of the duration of that contact. To model this feature of Bluetooth exposure notification systems and how it impacts vaccination prioritisation strategies, we present new theory in the next section that incorporates both the number and duration of contacts. This theory predicts that the total exposure time of contacts has a direct causal influence on the spread of infectious disease.

## Percolation on Weighted Networks

Many infectious diseases spread through close contact, and contact patterns in human populations display a high degree of heterogeneity [9, 22]. One successful approach to understanding the impact of heterogeneity is to model the spread of infectious disease as a percolation process on the network of contacts [29, 28, 38]. In the application of percolation theory to infectious diseases, the sum total of human contacts forms a network across the entire population. The vertices of this network are the individuals in the modelled population. An edge joins two individuals if they come into potentially infectious contact with one another in a typical time window matching the infectious period of the disease. The transmissibility of the infectious disease is described by the transmission probability, denoted *T*.

In a series of foundational works [32, 33], Newman developed analytical techniques based on probability generating functions. Through these techniques one can derive formulas for key epidemiological quantities in terms of summary statistics of the degree distribution of the contact network, that is, the distribution of the number of contacts made by individuals in the modelled population. In particular, Newman proved that the basic reproduction number, *R*_0_, depends on the transmission probability *T*, the average degree, and the variance of the degree distribution. As a corollary of these results, in particular the presence of the variance, we learn a fundamental insight: heterogeneity in contact patterns influences the spread of infectious disease.

The extent to which contact patterns differ between individuals goes beyond the number of contacts they have. Another key variable is the duration of each contact, as over the infectious period of the disease a typical individual will accumulate a large time in contact with a few regular contacts, and a much smaller amount of time with each of a potentially larger number of irregular contacts. Studies have shown that longer contact duration increases the transmission probability of COVID-19 [34]. Both the epidemiological significance of contact duration and the functionality of Bluetooth exposure notification systems imply the need to incorporate contact duration into the percolation framework.

We have generalized Newman’s analytic framework to understand percolation on *weighted networks* (see Methods). In our model, each edge, which represents a contact between individuals, is further equipped with a weight representing the duration. The transmission probability is assumed to depend on the duration of contact. This leads to replacing *T* with a distinct *T*_*w*_ for each weight.

We derive a formula showing that total exposure time of individuals is a driving factor in the spread of disease. As we discussed above, Bluetooth exposure notification apps measure each user’s total exposure time to other users of the app. Our model therefore predicts that total exposure time is an epidemiologically relevant quantity with a direct causal relationship to the reproduction number. Therefore, vaccine prioritisation via exposure notification apps is both plausible and potentially effective. To further examine the role of total exposure time in epidemic dynamics, in the next section we develop a quantitative framework for comparing prioritisation strategies.

Because of heterogeneities in contact patterns, vaccination of different individuals will have a varying impact on the rate of spread. In this regard, heterogeneity is a resource: preferential vaccination of those with greater total exposure time should lead to a greater reduction of disease spread from the same number of vaccine doses.

In order to compare different strategies we must further expand the percolation toolkit to incorporate vaccination. We model vaccination as a stochastic process which removes vertices from the original contact network. We assume that the strategies under consideration prioritise individuals according to the number and duration of their contacts, which is the case for strategies leveraging Bluetooth exposure notification apps. In the attached methods we derive formulas for the reproduction number on the residual network of susceptible individuals for a general vaccination strategy of this type.

## Assessing Vaccination Efficiency

To compare two vaccine prioritisation strategies it is not enough to consider the post-vaccination reproduction number without taking into account the number of individuals vaccinated by each strategy. This is especially true if one is concerned with how best to use a limited vaccine supply. To that end we introduce a novel measure to compare vaccine prioritisation strategies. We define the *efficiency* of a vaccination strategy to be:

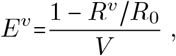

where *R*_*v*_ is the post-vaccination reproduction number and *V* is the fraction of the population that receives the vaccine. In other words, the efficiency of a strategy is the percentage decrease achieved in the reproduction number per percentage of the population vaccinated. It is important to note that since vaccination is modelled as a stochastic process, both *R*^*v*^ and *V*, and hence the efficiency, will vary across different realizations of the vaccination process.

The baseline strategy against which we compare prioritisation strategies is the uniform strategy, under which vaccines are distributed uniformly across the population to achieve a target vaccine coverage without taking into account any form of contact heterogeneity. The efficiency of the uniform strategy equals 1 since the expected reproduction number decreases linearly with the vaccine coverage from its original value. Strategies for which the expected efficiency is less than 1 are regarded as inefficient, since the same number of vaccines could have achieved a greater impact if they were allocated uniformly. On the other hand, strategies with efficiency greater than 1 are promising candidates for a significant impact. In the next section we introduce our exposure prioritisation strategy based on Bluetooth exposure notification apps whose efficiency is much greater than 1. We will refer to this vaccination strategy as “hot-spotting”, in reference to a fire-fighting practice that focuses on areas with intense fires [31].

## The Hot-spotting Strategy

Here we propose a “hot-spotting” vaccination strategy that prioritises app-using individuals for vaccination according to their total exposure time. The strategy operates without the central collection of any user data, as the prioritisation of each user is decided locally by their device.

The strategy depends on a parameter *β* encoding the probability of success in a weighted coin-flip. Each user performs a coin flip for each encounter stored in their encounter log over a fixed time period. Vaccines are prioritised to those who receive at least one success. Since the probability of obtaining at least one success on *n* weighted coin flips is 1 − (1− *β*)^*n*^, we see that users with a greater total exposure time have a greater probability of being prioritised for vaccination. Note that the extent to which high total exposure time individuals are prioritised can be increased by requiring a greater number of successes.

We simulate this strategy on a simulated network derived from a diary-based population contact survey (see Methods) [30]. In our model, individuals using the app form a subnetwork of the weighted network describing the population’s contact patterns. The number of entries in a user’s encounter log is their weighted degree in the app-user subnetwork. The results of the prioritisation strategy depend on the rate of app usage in the population, denoted *U*. In particular, the vaccine coverage *V* (proportion of individuals vaccinated) is bounded by the usage rate *U*.

In Figure 1 we show the simulation results for the efficiency as a function of vaccine coverage assuming a basic reproduction number of *R*_0_ = 1.5. The choice of basic reproduction number has little impact on the efficiency, as we show in the Supplementary Information by considering *R*_0_ = 2.2 as well. For very low vaccine coverage the expected efficiency is near 4 for all *U* values, in agreement with our theoretically derived value of 3.8. There is also a large variance in efficiencies due to small number statistics. As one expects from a strategy prioritising high exposure individuals, the efficiency decreases as vaccine coverage increases. This is because high exposure individuals are likely to be vaccinated first. Nonetheless, across the full range of vaccine coverage rates our proposed strategy is *one to four times more efficient than the uniform vaccination strategy*.

**Figure 1:**
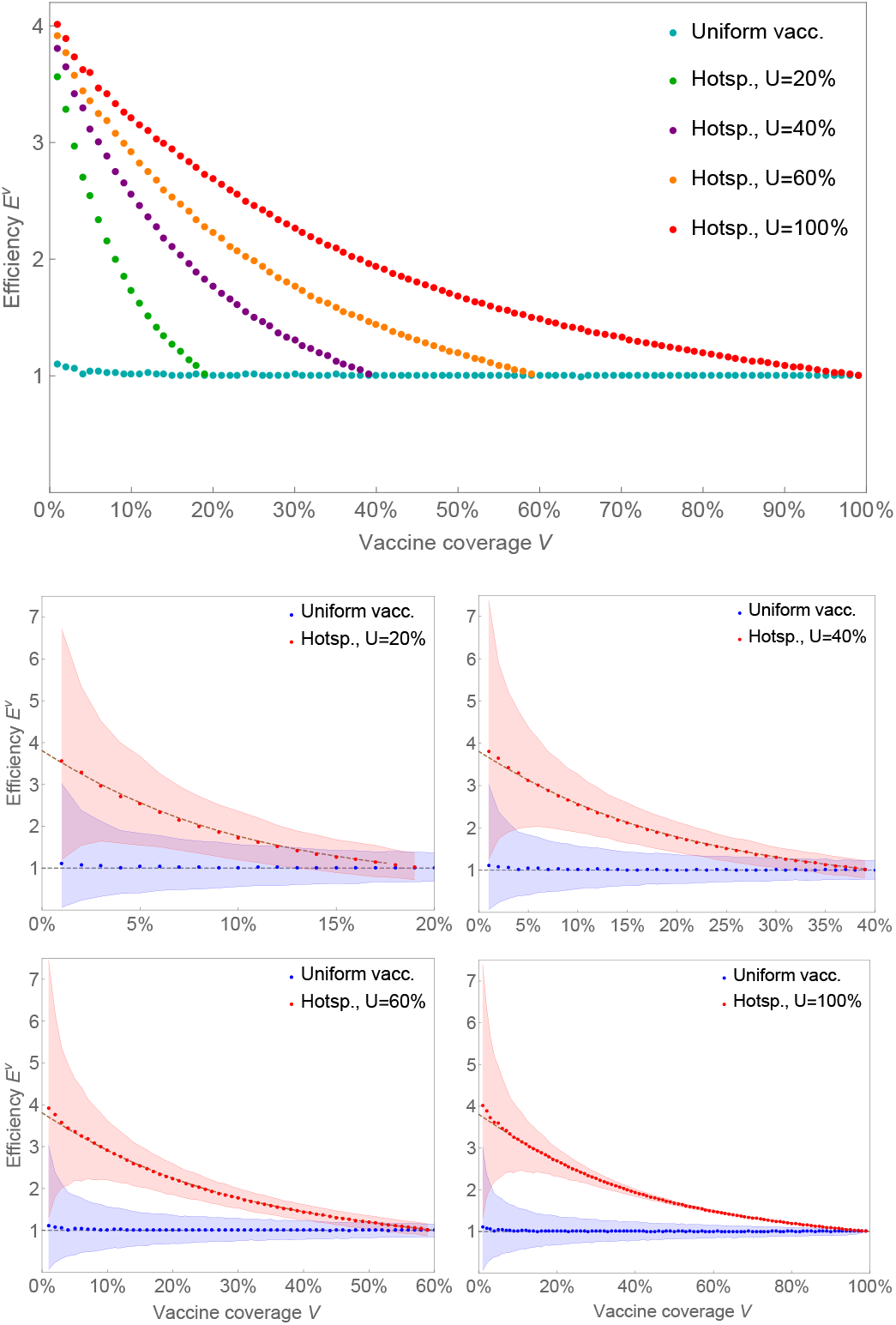
The efficiency for the hot-spotting strategy for *R*0 = 1.5 with various app usage rates are compared with the uniform vaccination in relation to vaccine coverage. The dots represent the mean efficiency obtained from 10,000 simulations for each dot. The shaded regions show the intervals that capture 90% of all simulation results. The dashed lines in the bottom figures show the expected efficiency obtained from the formula in Methods.

We observe that the expected efficiency curves for differing usage rates are simply the same curve rescaled. This implies that the important quantity to consider for assessing the efficiency is not the overall vaccine coverage *V* but rather the fraction of the app-using population which are vaccinated.

Figure 2 shows the simulation results for the post-vaccination reproduction number, assuming again an initial *R*_0_ = 1.5. The convexity of the curves is a witness to the superior efficiency of our proposed strategy. We see that as the fraction of the app-using population which are vaccinated increases the slope decreases, in alignment with the decreasing efficiency. The *greatest relative impact is achieved by vaccinating 20-40% of the app users*.

**Figure 2:**
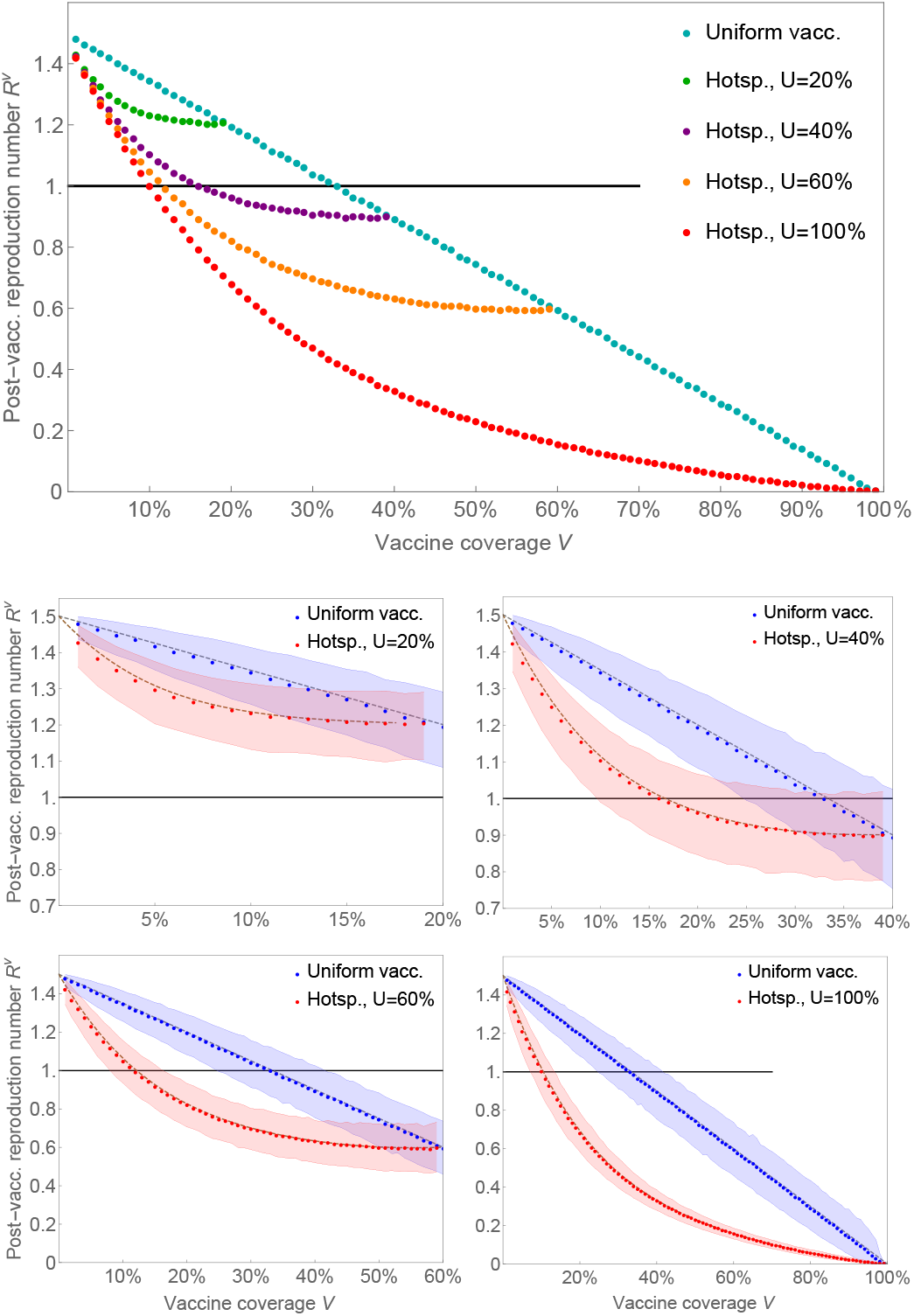
The post-vaccination reproduction numbers for the hot-spotting strategy for *R*_0_ = 1.5 with various app usage rates are compared with the uniform vaccination in relation to vaccine coverage. The dots represent the mean efficiency obtained from 10,000 simulations for each dot. The shaded regions show the intervals that capture 90% of all simulation results. The dashed lines in the bottom figures show the expected post-vaccination reproduction numbers obtained from the formula in Methods.

Finally, when app usage rates are high enough, hotspotting *achieves herd immunity with fewer than half as many doses*. In Figure 3 we show that the significantly lower herd immunity thresholds persist across a range of *R*_0_ values, and applies even when only 40% of the population uses the app.

**Figure 3:**
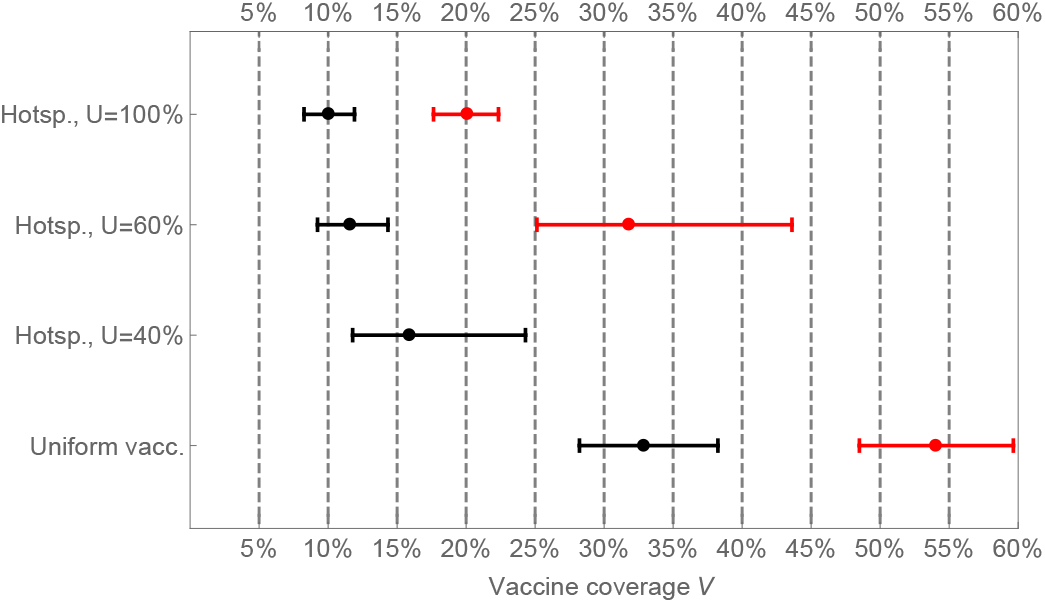
The necessary vaccine coverage to bring the reproduction number from its initial value *R*_0_ = 1.5 (black) or *R*_0_ = 2.2 (red) to the herd immunity threshold *R*^*v*^ = 1 is demonstrated for the uniform and hot-spotting strategy for various app usage rates. The dots represent the mean value from our simulations, whereas the intervals show the range that captures 1*σ* (68.27%) of all simulation results.

**Figure 4:**
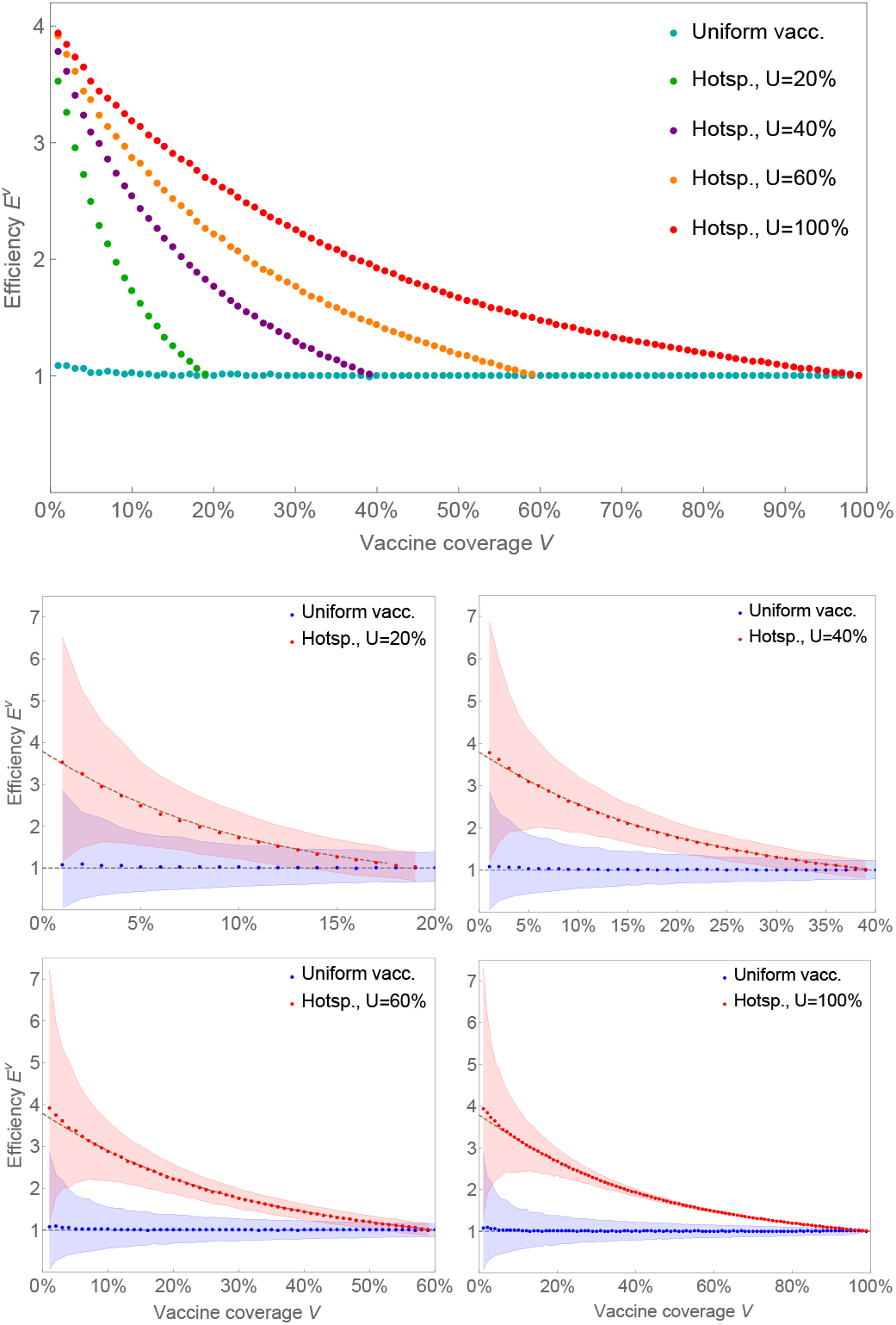
The efficiency for the hot-spotting strategy for *R*_0_ = 2.2 with various app usage rates are compared with the uniform vaccination in relation to vaccine coverage. The dots represent the mean efficiency obtained from 10,000 simulations for each dot. The shaded regions show the intervals that capture 90% of all simulation results. The dashed lines in the bottom figures show the expected efficiency obtained from the formula in Methods.

**Figure 5:**
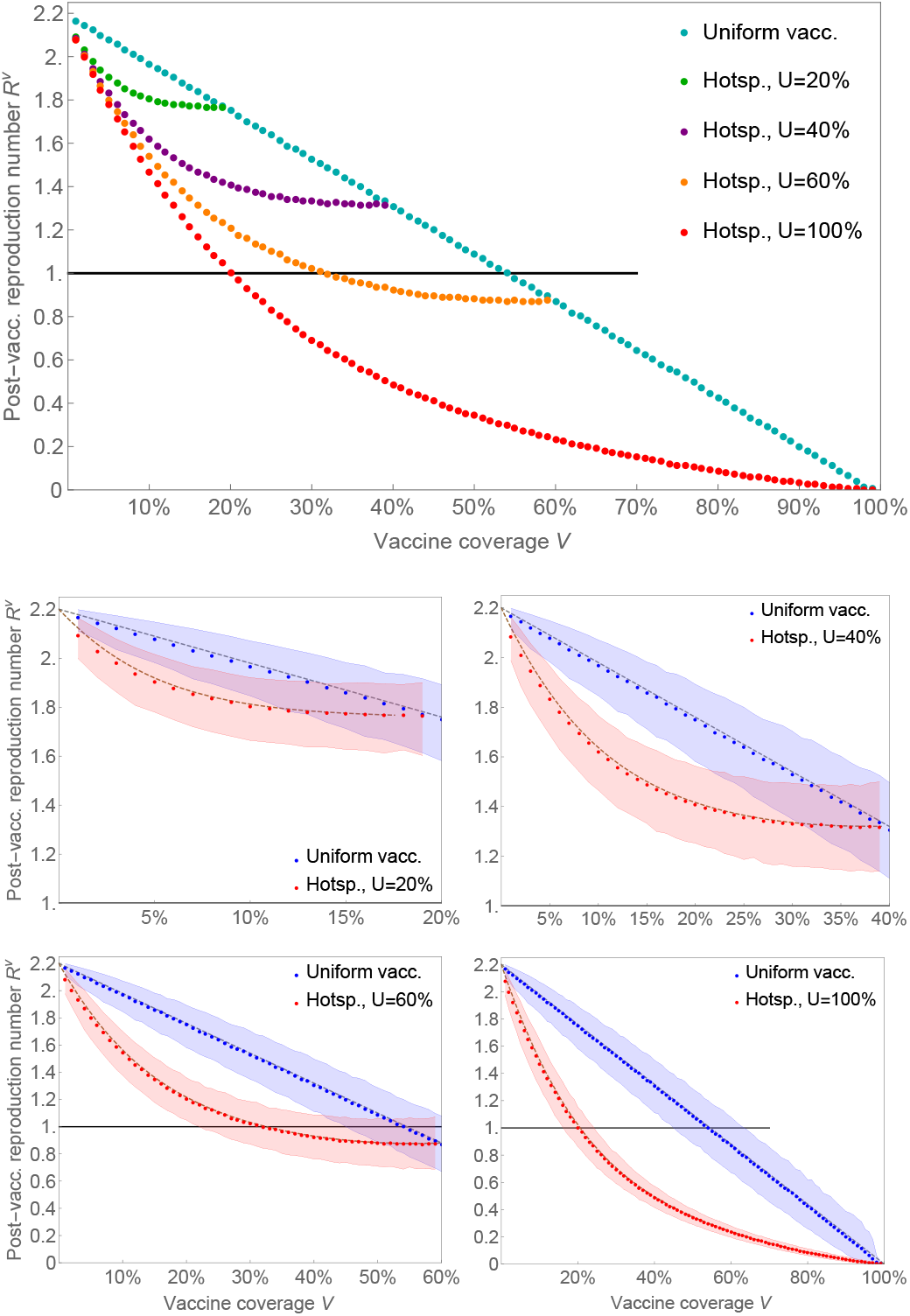
The post-vaccination reproduction numbers for the hot-spotting strategy for *R*_0_ = 2.2 with various app usage rates are compared with the uniform vaccination in relation to vaccine coverage. The dots represent the mean efficiency obtained from 10,000 simulations for each dot. The shaded regions show the intervals that capture 90% of all simulation results. The dashed lines in the bottom figures show the expected post-vaccination reproduction numbers obtained from the formula in Methods.

## Concluding Remarks

Implementing our proposal in practice is surprisingly easy on the technical front, although it requires the participation of key players. The exposure prioritisation scheme itself is straightforward and requires no modifications of the core Bluetooth exposure notification framework. Since the Google/Apple system is implemented at the operating system level it is Google and Apple who must incorporate this new functionality. Public health authorities who wish to use hot-spotting must also ensure that their vaccine allocation systems are coordinated with the functionality of the app. Our proposal is, moreover, not strictly limited to If *R*_0_ = 2.2 and *U* = 40%, vaccinating only the app users would not be enough to reach herd immunity. digital contact tracing solutions based on Bluetooth. The hot-spotting strategy can be added on to any technology which creates an encounter log for each user.

As for any vaccine prioritisation strategy, hot-spotting comes with ethical concerns, including questions of access, equity, and vaccine program objectives. In addition, if not carefully integrated as part of a holistic approach to public health measures, a naive implementation of this approach could lead to perverse incentives for people to increase their exposure or circumvent the system. We are proposing hot-spotting as part of a broader approach that addresses equity and is implemented in a way that pre-empts harmful behavioural responses [40].

An important limitation of our approach is the underlying data used to construct the model network, which was collected before the pandemic [30]. Moreover, in both our theoretical work and our simulations we have assumed that there is no distinction between a potentially infectious contact and the contacts detected by Bluetooth exposure notification apps. Finally, our modelling does not predict the time evolution of the epidemic in response to vaccination.

These simplifying assumptions could be relaxed in future work with more detailed agent-based simulations that test the generality of the new theory we have introduced. However, we note that a diverse collection of previous literature already finds that prioritising individuals based on their number of contacts can be highly effective [35, 19, 16, 14]. We have built upon this literature by (1) proposing a measure *E*^*v*^ of the relative efficiency of different strategies, (2) showing that existing COVID-19 digital contact tracing technology allows the measurement of epidemiologically important quantities without violating privacy, (3) proposing a new vaccination strategy based on these measures that is technically feasible to implement, and (4) introducing novel analytical techniques and simulations that (5) demonstrate how hot-spotting is dramatically more efficient than uniform allocation. We conclude that hot-spotting could enable public health authorities to greatly reduce the social and economic costs of COVID-19 until vaccine supply catches up with demand.

## Data Availability

All source files for the simulations are available upon request.

## Acknowledgements

We acknowledge support from the National Sciences and Engineering Research Council of Canada (NSERC), NSERC COVID-19 Alliance Grant (MDP, MA, CTB) and Ontario Ministry of Colleges and Universities COVID-19 Rapid Response Grant (MA, CTB).

Patent pending, United States Provisional Application No. 63/122,587.

## Methods

We model the spread of infectious disease in a population as a percolation process on the network of contacts. Each vertex in this network represents an individual person and each edge links two people who come into contact. Our particular approach is a generalization of Newman’s *probability generating function* framework [33] to weighted networks.

### Percolation Theory

The key mathematical object in the original formalism is the probability generating function of the degree^1^ distribution,

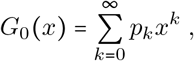

where *p*_*k*_ is the fraction of vertices in the network having degree *k*. This function is useful for writing analytic expressions for moments of the distribution; for example, the average degree is given by 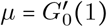.

An infectious disease spreads in the network through occupied edges. In the original model, each edge has a uniform probability *T* for being occupied. The basic reproduction number^2^ *R*_0_ corresponds to *the expected number of other occupied edges attached to a vertex at the end of a random occupied edge*. This number is given by

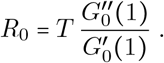

In terms of the mean *µ* and the standard deviation *σ* of the degree distribution, this equation is equivalent to

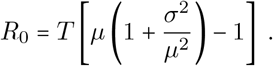

### Weighted Percolation

In order to model vaccine prioritisation strategies based on exposure notification apps it is necessary to incorporate contact duration into the percolation model. To do this we consider the contact network to be weighted: each edge is further equipped with a weight *w*. The transmission probability along an edge for the percolation process is assumed to depend on that edge’s weight, so that the single parameter *T* is replaced with a distinct *T*_*w*_ for each weight *w*.

For our purposes in this paper the weight represents the number of time-steps over which the contact took place. However, the weights could represent any factor which influences the transmission probability along an edge, such as the nature of the contact, the setting, or the presence or absence of PPE. Unless stated otherwise, our analytical results hold for this general interpretation of weights. Given that this is quite a flexible formalism it can be applied to a broad range of modelling tasks.

For weights representing contact duration there is a natural candidate for the transmission probabilities *T*_*w*_. If one assumes that in each time-step there is an independent probability *T*_1_ of transmission, the transmission probability after *w* time-steps is

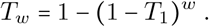

Once again, *T*_*w*_ is the probability for a weight-*w* edge to be occupied. The infection spreads through occupied edges.

In a weighted network we represent the configuration of edges around each vertex by a *generalized degree*, denoted ***k***. This is a vector having an entry for each of the possible weights appearing in the network. For a vertex of generalized degree ***k***, the entry corresponding to weight *w*, denoted *k*_*w*_, is the number of contacts having weight *w*. There are two important quantities we can extract from ***k***: First, the degree of a vertex, that is, the number of distinct contacts, is simply ∑ _*w*_ *k*_*w*_. Second, the *weighted degree* of a vertex, that is, the sum of the weights on each edge, is ∑ _*w*_ *wk*_*w*_. When *w* represents contact duration, the weighted degree is the total exposure time of that individual to others.

To generalize Newman’s results to the weighted setting we introduce a *multivariable generating function* for the distribution of generalized degrees. There is one variable *y*_*w*_ for each weight *w* appearing in the network, and we write ***y*** for the vector of these variables. Let *q*_***k***_ denote the fraction of vertices in the network having generalized degree ***k***. Then the multivariable generating function of the network is

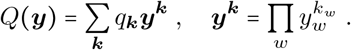

The generating function *G*_0_ (*x*) for the degree distribution in a weighted network is related to this function by *G*_0_ (*x*) = *Q* (**1***x*), where **1** is a vector with each entry being 1.

In order to derive the basic reproduction number, we need the distribution of *occupied degree*, i.e., the number of occupied edges attached to a vertex. This distribution is given by

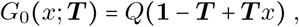

where ***T*** is the vector composed of the transmission probabilities *T*_*w*_. Furthermore, we introduce

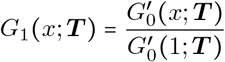

as the probability distribution of the number of other occupied edges attached to a vertex reached by following a random occupied edge (cf. [33]). By definition, the basic reproduction number *R*_0_ is precisely the mean of this final distribution, i.e.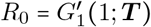.

One can express *R*_0_ directly in terms of the generating function *Q*(***y***). This is accomplished by introducing the differential operator ∇ _***T***_, defined as

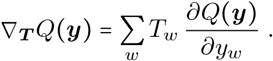

Then, we Find

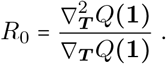

Alternatively, this result can be rewritten in a form which is more amenable to computation,

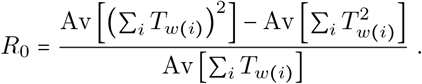

In this equation, the sums ∑ _*i*_ are taken over the contacts of each vertex, with *w*(*i*) being the weight of that contact, and the averages Av [·] are taken over all vertices in the network. Note that this formula holds regardless of the interpretation of the weights as contact duration.

### Total Exposure Time

Let us now focus on the case when the weights represent contact duration and *T*_*w*_ =1 −.(1 − *T*_1_)^*w*^. In this case, the weighted degree of a vertex is the total exposure time of that individual to others, measured in discrete time steps. To better understand the impact of the total exposure time on the reproduction number, consider the approximation *T*_*w*_ ≈*wT*_1_, which is valid when *wT*_1_<<1 for all *w*. Under this approximation, we find the heuristic formula

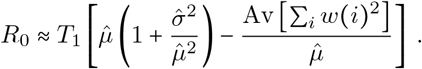

Here, 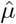 and 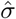 are the mean and the standard deviation of the total exposure time. Our model therefore predicts that the total exposure time has a direct influence on the spread of disease.

### Vaccination on Contact Networks

We model vaccination as a stochastic process wherein a vertex of generalized degree ***k*** has a probability *v* (***k***) of being vaccinated. Vaccination modifies the original contact network by *removing vaccinated vertices*, as we assume that a vaccinated individual can neither become infected nor infect others.

We introduce the *post-vaccination reproduction number R*^*v*^ as the reproduction number in the non-vaccinated sub-population after a vaccination process. Since vaccination is modelled as a stochastic process, *R*^*v*^ is a random variable.

The formulas for the basic reproduction number *R*_0_ can be adapted to give the expected post-vaccination reproduction number. This adaptation requires two changes: Firstly, some vertices are removed themselves. Each vertex of generalized degree ***k*** has a probability (1 − *v*(***k***)) of remaining in the residual network. Secondly, some contacts of the remaining vertices are removed. A weight-*w* contact has the probability

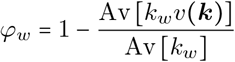

of remaining non-vaccinated. Hence, the expected post-vaccination reproduction rate is given by

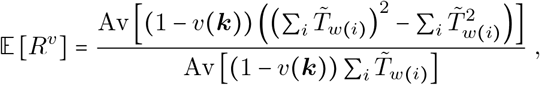

Where 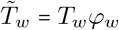, and the averages Av[…] on the right-hand side run through all vertices in the network.

### Efficiency of Vaccine Prioritisation

In general, one expects that prioritising the vaccination of individuals with greater exposure to others achieves a greater reduction in *R*^*v*^ than vaccinating the same number of less-connected individuals. This translates to slowing the infection more effectively and a reduction in the number of infections. Vaccine prioritisation becomes particularly important when only a small supply of vaccines is available. In that situation, adopting a strategy that prioritises highly connected individuals for vaccination can save the lives of more people.

To compare different prioritisation strategies we are concerned with not only how much they reduce the spread of disease, but also how many vaccines it takes for that outcome. To that end, we introduce a novel measure of prioritisation efficiency,

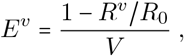

where *V* is the fraction of the population which receives the vaccine.

In a stochastic process of vaccination, *V, R*^*v*^ and *E*^*v*^ are all random variables. In particular, the vaccine coverage rate *V* has the expected value 𝔼 [*V*] = Av [*v* (***k)***] and the variance Var 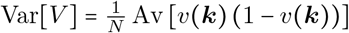, where *N* is the size of the population. Therefore, for computing the expectation value of the efficiency *E*^*v*^ in a large population, we may neglect the variance in *V* and treat it as a fixed number at its mean value. Hence, we can write

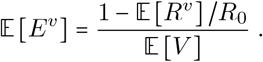

For a uniform vaccination probability *v* without any prioritisation, one would have 𝔼 [*R* ^*v*^] = (1 − *v*) *R*_0_ and 𝔼 [*E*^*v*^] = 1. Through a realistic vaccine prioritisation strategy, it is possible to achieve a significantly higher efficiency, while the exact numbers depend on the underlying network.

### The Hot-spotting Strategy

We propose a *hot-spotting strategy* that can be implemented simply on the existing Bluetooth based exposure notification apps. This strategy is based on a Bernoulli process for each exposure registered in the app with a chance of *β* for success. The app users with at least one success are given the priority for vaccination. Hence, if an app user has a total exposure number *n* in the app, they will have a chance of 1 − (1 − *β*) ^*n*^ for being selected for prioritised vaccination in this strategy.

In order to estimate the impact and efficiency of our hot-spotting strategy on the whole population, we must take into account that not every person in the population uses the app in question. To this end, we assume that there is an app usage rate of *U* and that the app users are homogeneously distributed in the population. We also assume that there is no preferential attachment between app users. Then, for a random individual with the contact structure ***k***, which is not necessarily registered in the app network, the probability for them being an app user and also being selected for prioritised vaccination is given by

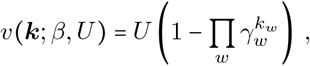

where

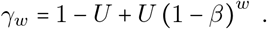

### Model Network

The well-known POLYMOD study surveyed over 7000 individuals across 8 European countries [30]. Respondents kept a log of all contacts made on a single day noting, among other features, how long the contact lasted and how frequently that contact is made. In the language of time-weighted networks, the inclusion of duration data means that the survey responses sample from the generalized degree distribution of the daily contact network.

To model COVID-19, the network should capture the contacts made over the typical infectious period of the disease. This varies significantly between patients, with one study estimating a median of 8 days after the onset of symptoms [42]. For simplicity we choose a period of 14 days, as it aligns well with the frequency responses in the survey.

Using the daily contact data we must generate samples from the generalized degree distribution of the *fortnightly contact network*. We accomplish this by a bootstrapping technique. More precisely, for each contact recorded, respondents chose between 5 options concerning both the duration and the frequency of that contact as shown in Table 2. We can therefore represent each respondent’s contacts on that day in a 5 × 5 matrix whose (*i, j*)entry is the number of contacts recorded with frequency key *i* and duration key *j*. For respondent *n* we denote this matrix by *D*_*n*_.

**Table 1:** Exposure notification app download rates in selected countries based on official sources.

| Country | Downloads per 100 people |
| --- | --- |
| Canada [2] | 20 |
| Germany [8] | 29 |
| Italy [6] | 17 |
| New Zealand [1] | 49 |
| Scotland [5] | 27 |
| Singapore [7] | 56 |

**Table 2:**
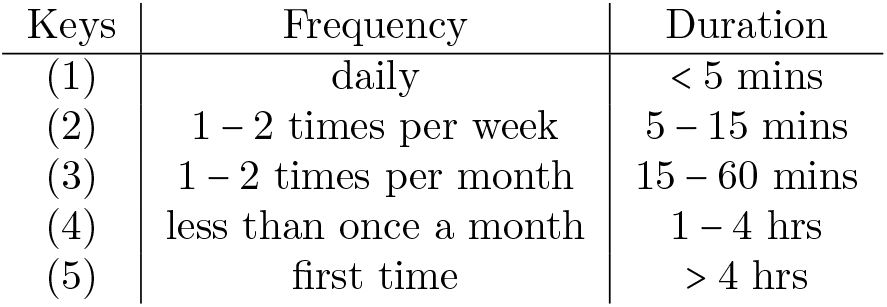
Possible responses on POLYMOD survey concerning frequency and duration of contact.

Our goal is to extrapolate fortnightly contact matrices *F*_*n*_ from the daily contact matrices *D*_*n*_. In our boot-strapping procedure, it is important that we distinguish between daily repeating contacts, which have frequency key (1), and *infrequent contacts*, which have frequency keys (2)-(5). We denote by *I*_*n*_ the matrix of infrequent contacts for respondent *n*. The matrix *I*_*n*_ is obtained from the same respondent’s *D*_*n*_ by setting the first row of daily repeating contacts to 0.

The matrices *F*_*n*_ are created by sampling from the daily contact matrices. Each sample is generated as follows:

1. Sample 14 respondents, *n*_1_, *…, n*_14_.
2. Set 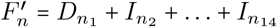.
3. Produce *F*_*n*_ from 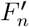 by dividing each entry in the second row by 3 and rounding it to the nearest integer.

In words, we sample 14 daily contact logs from the survey. Then, we add together the 1-day duration-frequency matrices for each of the samples, excluding the daily repeating contacts from all but the first sample. Then, assuming that a frequency-key-(2) contact is seen 3 times in a fortnight, we divide the second row by three to account for repeated counting of the same contact.

Finally, we assign to each type of contact in the matrix *F*_*n*_ a certain weight. Each unit of weight is approximately 10 minutes of contact time, which corresponds to the token exchange rate of the exposure notification apps. We chose these weights to be given as in the following weight matrix:

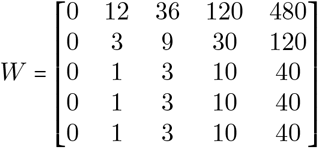

Hence, we obtain a list of generalized degrees sampled from a virtual weighted contact network, in which the set of possible weights is {1, 3, 9, 10, 12, 30, 36, 40, 120, 480}.

### Simulation

The simulations presented in the main text are based on 500 samples of fortnightly generalized degree distributions generated through the bootstrapping procedure described above. We interpret these 500 samples as an *observed cohort* of 500 individuals within a much larger population. As such, the recorded contacts are assumed to lie outside the observed cohort.

For the chosen data of 500 samples, we fix the unit transmission probability at *T*_1_=0.000375 to get the basic reproduction number *R*_0_= 1.501. We select a set of parameters *β* for the vaccination probability function *v (****k***; *β, U)* of the hot-spotting strategy (and a set of parameters *v* for the uniform strategy) to target a homogeneous distribution of the vaccinate rate *V* in the outcome over its range of possible values. 10,000 simulations are performed for each selected parameter. This accounts to a total of 990,000 simulations for the strategies with *U =* 100%. For the strategies with *U =* 20%, 40%, and 60%, we ran a total of 190,000, 390,000, and 590,000 simulations, respectively.

Each simulation consists of four steps:

1. The vaccination probabilities *v* (***k***; *β*,) for each individual, and the probabilities *ϕ*_*w*_ for each weight are calculated for the given parameters.
2. For each component *k*_*w*_ of the generalized degree of each individual, a binomial process is performed with the probability *ϕ*_*w*_. The outcome of this process replaces *k*_*w*_ as the residual contact number.
3. For each individual, a Bernoulli trial is performed with the probability *v* (***k***; *β, U*). If the outcome is a success, the individual is removed from the list with all their contacts, otherwise they remain.
4. The base reproduction number is computed in the residual list and saved as the post-vaccination reproduction number *R*^*v*^. The vaccine coverage *V* is deduced from the length of the residual list. The efficiency *E*^*v*^ is calculated from *R*^*v*^ and *V*.

## Supplementary Information

1 The degree of a vertex is the number of edges attached to it.

2 In heterogeneous populations, *R*_0_ is defined as “the expected number of secondary cases produced, in a completely susceptible population, by a typical infected individual during its entire period of infectiousness” [15].

